# A panel-agnostic strategy ‘HiPPo’ improves diagnostic efficiency in the UK Genome Medicine Service

**DOI:** 10.1101/2023.01.31.23285025

**Authors:** Eleanor G. Seaby, N. Simon Thomas, David Hunt, Diana Baralle, Heidi L. Rehm, Anne O’Donnell-Luria, Sarah Ennis

**Affiliations:** Human Development and Health, Faculty of Medicine, University Hospital Southampton, Southampton, Hampshire, SO16 6YD, UK; Program in Medical and Population Genetics, Broad Institute of MIT and Harvard, Cambridge, MA 02142, USA; Division of Genetics and Genomics, Boston Children’s Hospital, Boston, MA 02115, USA; Paediatric Infectious Diseases, Imperial College London, London, W2 1NY, UK; Wessex Regional Genomics Laboratory, Salisbury NHS Foundation Trust, Salisbury, SP2 8BJ, UK; Center for Genomic Medicine, Massachusetts General Hospital, Boston, MA 02114, USA

## Abstract

Genome sequencing is now available as a clinical test on the National Health Service (NHS) through the Genome Medicine Service (GMS). The GMS have set out an analytical strategy that predominantly filters genome data on a pre-selected gene panel(s). Whilst this approach reduces the number of variants requiring assessment by reporting laboratories, pathogenic variants outside of the gene panel applied may be missed, and candidate variants in novel genes are largely ignored.

This study sought to compare a research exome analysis to an independent clinical genome analysis performed through the NHS for the same group of patients. When analysing the exome data, we applied a panel agnostic approach filtering for variants with **<u>Hi</u>**gh **<u>P</u>**athogenic **<u>Po</u>**tential (HiPPo) using ClinVar, allele frequency, and *in silico* prediction tools. We then compared this gene agnostic analysis to the panel-based approach as applied by the GMS to genome data. Later we restricted HiPPo variants to a panel of the Gene Curation Coalition (GenCC) morbid genes and compared the diagnostic yield with the variants filtered using the GMS strategy.

24 patients from 8 families underwent parallel research exome sequencing and GMS genome sequencing. HiPPo analysis applied to research exome data identified a similar number of variants as the gene panel-based approach applied by the GMS. GMS clinical genome analysis identified and returned 2 pathogenic variants and 3 variants of uncertain significance. HiPPo research exome analysis identified the same variants plus an additional pathogenic variant and a further 3 *de novo* variants of uncertain significance in novel genes, where case series and functional studies are underway. When HiPPo was restricted to GenCC disease genes (strong or definitive), the same pathogenic variants were identified yet statistically fewer variants required assessment to identify more diagnostic variants than reported by the GMS genome strategy. This gave a diagnostic rate per variant assessed of 20% for HiPPo restricted to GenCC versus 3% for the GMS panel-based approach. With plans to sequence 5 million more NHS patients, strategies are needed to optimise the full potential of genome data beyond gene panels whilst minimising the burden of variants that require clinical assessment.

## Introduction

Genome sequencing is now available as a diagnostic test on the National Health Service (NHS) in the UK, offered through their Genome Medicine Service (GMS). With the cost of genome sequencing becoming ever competitive, genome sequencing is beginning to supersede exome sequencing in some institutes, including in the NHS.(1) However, one of the challenges in diagnosing patients with rare disease is the expanded scope of analysis and need to correlate results with phenotype.(2) Genome sequencing produces 3-4 million variants per individual; therefore, strategies to reduce noise and focus on the most salient regions of DNA have been adopted, including use of virtual gene panels.(3, 4) For the NHS, this is their primary analytical strategy, meaning that despite sequencing and storing an entire genome, only a fraction of the genome is actually analysed. Consequently, this risks missing pathogenic variants that would have been identified if more regions of the genome had been assessed.

All that said, there remains a trade-off between utilising the breadth of sequencing data available (such as for a genome) and the number of variants that require assessment by clinical laboratories. Filtering is necessary to reduce the number of variants identified to a manageable number that NHS laboratories can analyse, classify with respect to pathogenicity, and interpret with respect to causality of the patient’s symptoms in a reasonable and acceptable timeframe.

The GMS, which primarily sequences trios, adopts a workflow similar to that used in the 100,000 Genomes Project, which predated the GMS.(1, 5) First the data are filtered by inheritance pattern(s), data quality, and allele frequency. Following this, the remaining variants are filtered by a gene panel(s) selected by the clinician when the test is ordered. Short variants and copy number variants (CNVs) overlapping the gene panel are returned for analysis (“Tiered variants”). The only variants mandated to be assessed outside of the gene panel(s) are ‘gene agnostic variants’ comprising *de novo* coding variants and Exomiser(6) top 3 ranked variants which are not filtered on quality (**Figure 1**).

**Figure 1.**
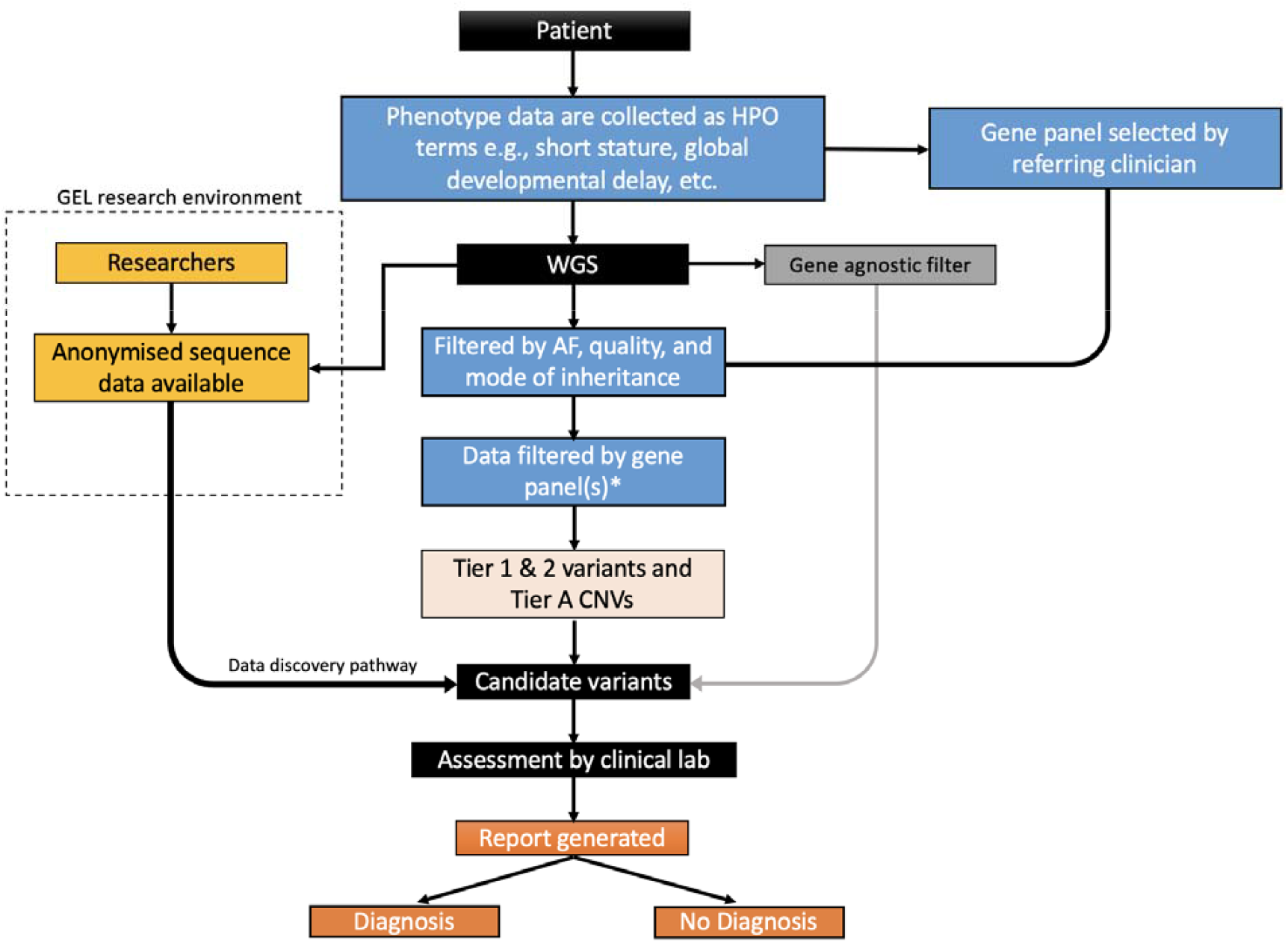
Genome Medicine Service workflow for genome sequencing on the NHS Tier 1 variants are defined as predicted loss-of-function variants or de novo variants in a green gene on the PanelApp gene panel(s) applied. Tier 2 variants are defined as coding variants +/- 8bp (excluding synonymous) on any transcript in the panel applied. Synonymous variants affecting splicing are ignored. The gene agnostic filter includes top 3 Exomiser rank variant with score of ≥0.95 and any de novo (coding) variant. Tier A is defined by a CNV (>10KB) overlapping a ClinGen curated pathogenic region relevant to a panel applied or a CNV overlapping with a green gene in the panel applied. Anonymised sequencing data are available for some patients in the Genomics England (GEL) Research Environment. *Gene panels are selected using GEL PanelApp by the referring clinician.

In contrast to genome sequencing, exome sequencing targets only coding regions of DNA. However, most variants filtered in the GMS strategy (Tier 1, Tier 2, and gene agnostic variants) would be captured by an exome. Given the method limitations of exome sequencing, genomes offer better coverage (even for coding regions) than exomes do and are far superior for identifying CNVs and other structural variants.(7) All that said, genome data are costly to store and process computationally and this should be considered alongside the benefits to having access to non-coding data, particularly if those data are mostly ignored.

Panel based approaches that restrict analyses to clinically relevant genes clearly have merit, yet 26% of diagnoses made through the 100,000 Genomes Project were not on the original gene panel applied.(8) Therefore, complementary approaches that look beyond gene panels are warranted. However, this must be balanced with the potential of increasing the number of variants that require assessment by reporting laboratories. Currently the GMS assess every variant that is in a ‘green’ gene in the PanelApp(4) gene panel applied, regardless of *in silico* predictions. Metrics such as CADD(9), REVEL(10), and SpliceAI(11) can help reduce noise, facilitating the assessment of variants across a wider spectrum of genes without too additional burden. We sought to exploit this principle by adopting a panel agnostic approach that filters variants on **<u>Hi</u>**gh **<u>P</u>**athogenic **<u>Po</u>**tential (HiPPo) across the exome by utilising *in silico* prediction scores, allele frequency, and ClinVar(12) (**Figure 2**).

**Figure 2.**
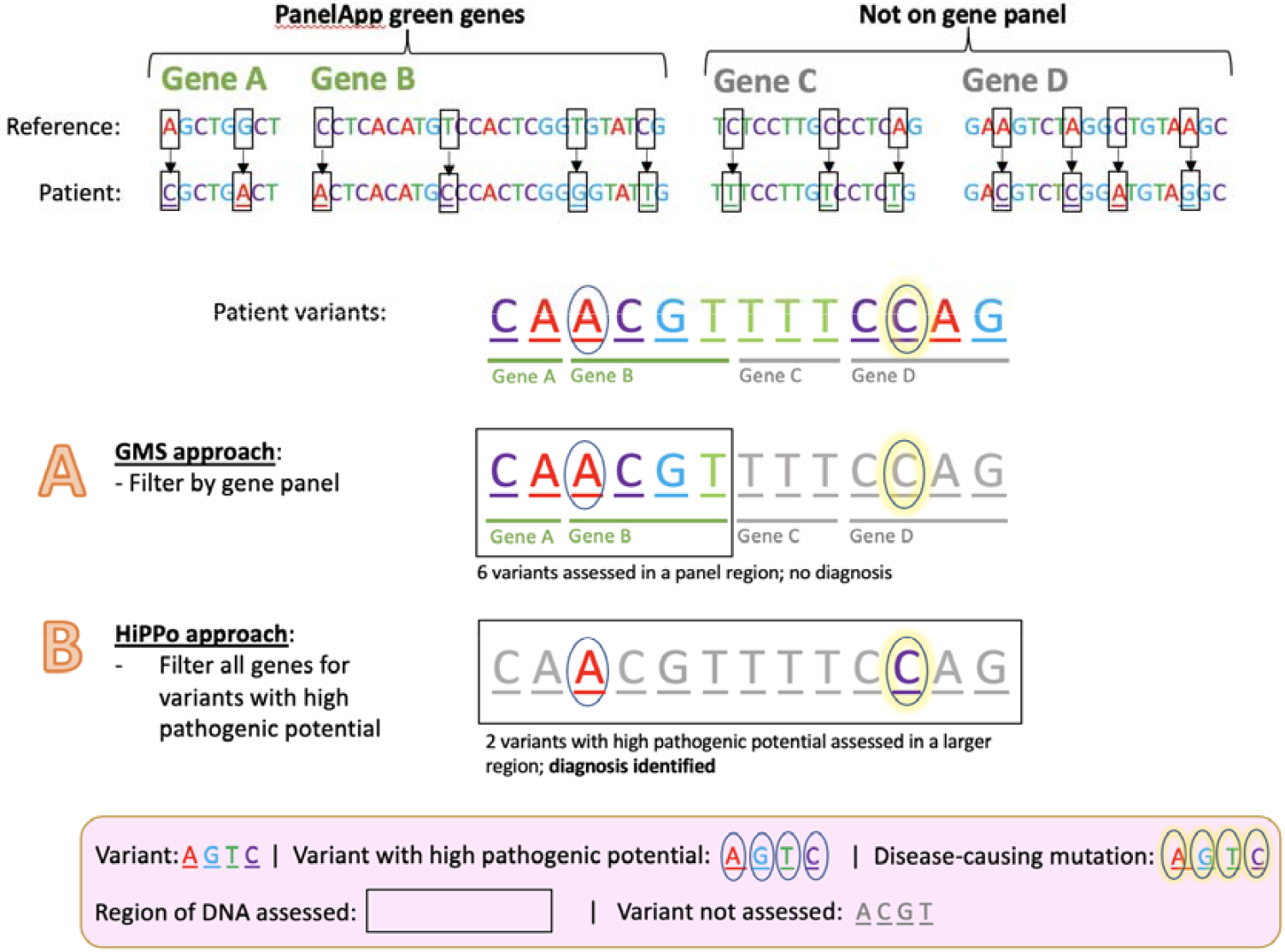
A proposed method for improving diagnostic yield and efficiency Comparison of the current NHS approach versus our proposed method HiPPo. Variants are identified by comparing a patient’s DNA against a human genome reference. In this example, there is a pathogenic variant (yellow highlighted circle) within the identified list of variants. To minimise the number of variants assessed, the NHS has adopted a method (**A**) that looks in small regions of the DNA (a panel of genes) and assesses the variants within that region. If the causal variant is in a region of the DNA not assessed, then the diagnosis is missed. Our revised approach captures a larger region of DNA (including all genes), but only looks at variants predicted to be damaging or submitted as P/LP to ClinVar (black circle). As a result, a larger area of DNA is assessed, whilst assessing fewer variants overall. This aims to result in a higher diagnostic rate per number of variants assessed, despite analysing a larger region of the genome than typically applied in a gene panel.

This study compares the analysis of exome sequencing data performed in a research setting with genome sequencing performed on the same patients through the GMS in a clinical setting. We adopt a gene-agnostic approach, HiPPo, and compare the diagnostic yield of this approach with the strategy applied by the GMS. We aim to improve upon both the efficiency and diagnostic rates of current GMS standards, whilst trying to minimise the number of variants requiring assessment by clinical laboratories.

## Methods

### Recruitment and patient demographics

Clinical Geneticists at University Hospital Southampton were invited to recruit patients and families with suspected monogenic disease to a research study ‘*Use of NGS technologies for resolving clinical phenotypes*’ (IRAS: 212945; REC: 17/YH/0069). Recruited individuals were eligible for a research exome through the Center for Mendelian Genomics(13) at the Broad Institute.

Twenty-seven individuals from nine families recruited to the research exome study were also recruited for genome sequencing on the NHS through the GMS, facilitating a parallel comparison study (**Figure 3**), providing an opportunity to evaluate these two sequencing and analysis strategies. All participants consented for their data to be shared.

**Figure 3.**
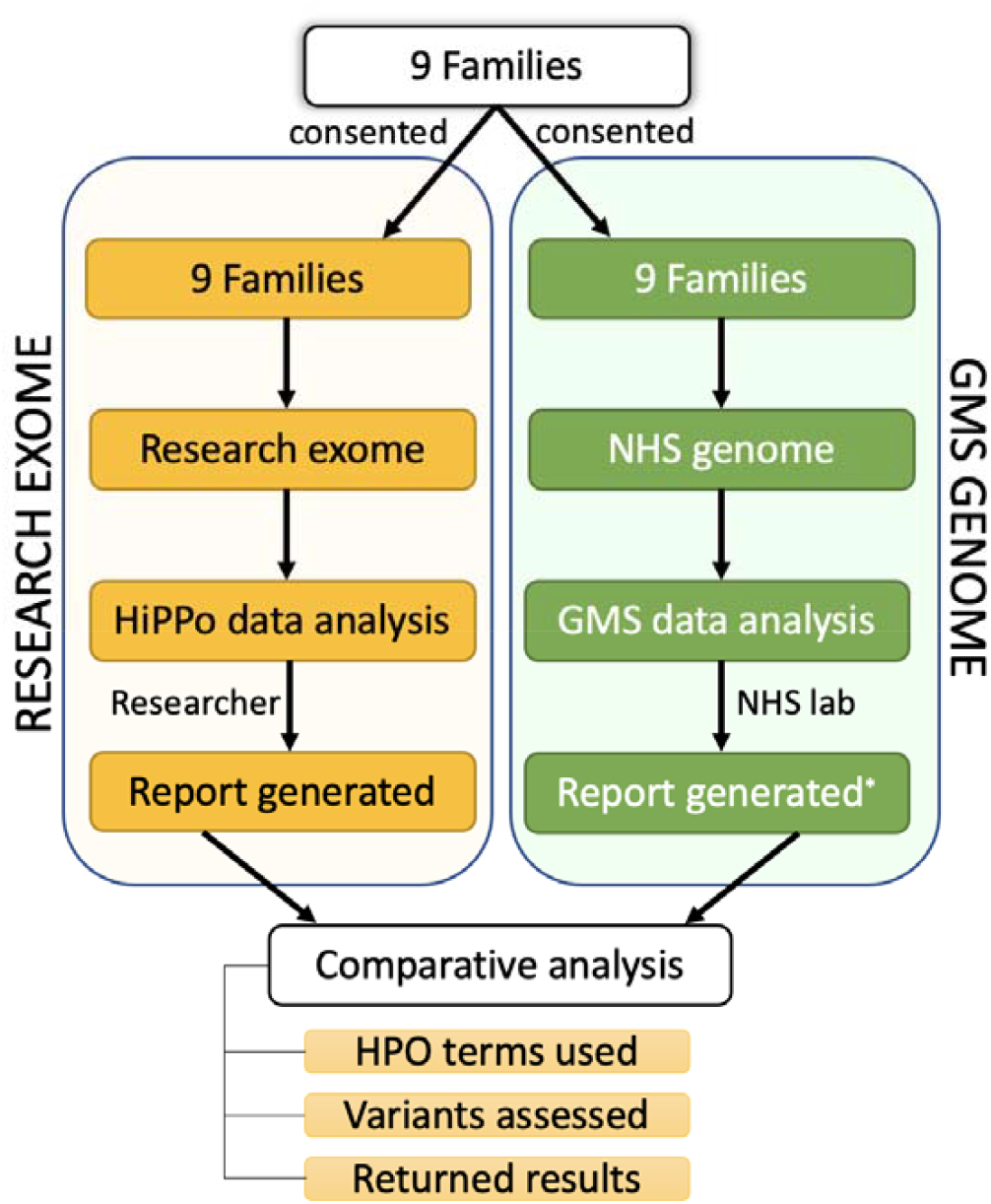
Overview of patient recruitment and analysis 9 families were recruited for parallel GMS clinical genome sequencing and research exome sequencing. Different data analysis strategies were applied to the exome (HiPPo) vs genome sequencing data (adopting a panel-based strategy as outlined by the GMS). Variants reported were compared between analysis strategies including the Human Phenotype Ontology (HPO) terms used, number of variants assessed, and results reported. *8 families with complete reports

For the research exome study, patient phenotypes were extracted by a single researcher from the clinical notes and recorded as Human Phenotype Ontology (HPO) terms in a manually encrypted database. The patient’s clinician also separately recorded HPO terms when requesting the GMS genome sequencing test. Both clinician and researcher were blinded to each other’s curated HPO terms. The family structures of the 9 families (8 trios and a quad), individual IDs, and phenotypes are described in **Table 1**.

**Table 1.**
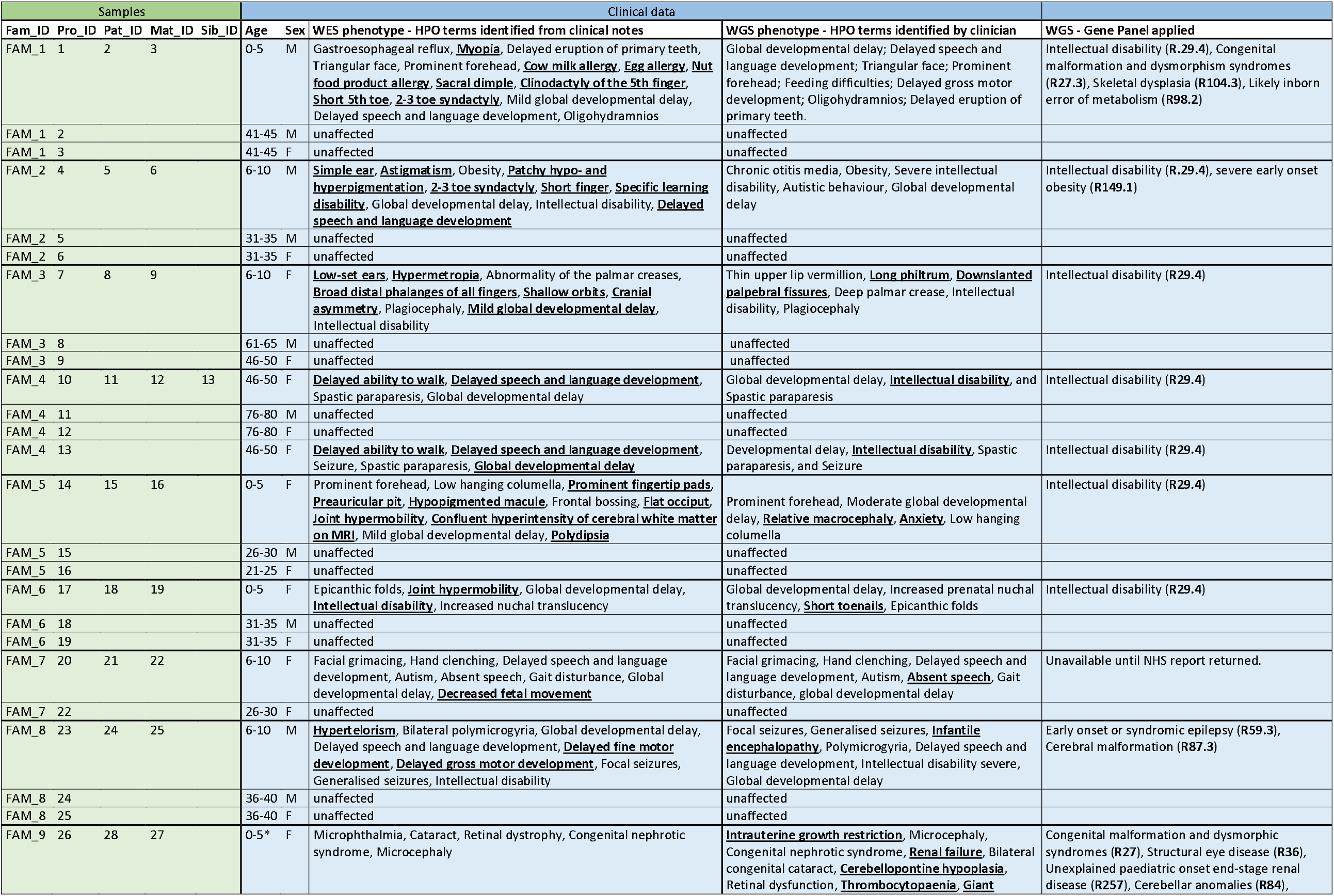
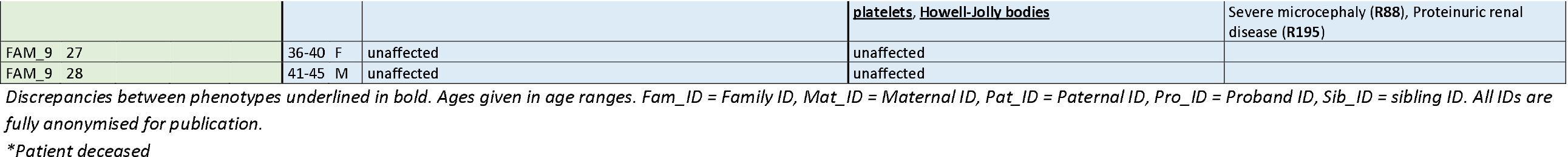
Samples and phenotypes of patients recruited for a parallel research exome and NHS genome

#### Research exome sequencing and pipeline

Following quality control of the DNA from the 27 samples, the mother (FAM_4_12) in the family comprising a quad of parents and monozygotic twins (FAM_4) had insufficient DNA quality, and we were unable to obtain a repeat sample in time for inclusion in the research exome portion of this study. However, this participant had genome sequencing through the GMS. In the GMS, quads are sequenced as two separate trios, therefore family FAM_4 was exome sequenced without maternal data (father, twin A, twin B) for the research portion of the study, but was genome sequenced through the GMS as two separate trios (mother, father, twin A) and (mother, father, twin B).

A total of 26 samples from 9 families met quality standards necessary for research exome sequencing at the Broad Institute (**Supplementary Data Table 1**). Libraries from DNA samples were created with an Illumina exome capture (37 Mb target) and sequenced on a NovaSeq 6000 machine using the NovaSeq XP workflow to cover >85% of targets at >20x, comparable to ∼55x mean coverage. The samples underwent QC as previously described and were processed through the GATK best practices pipeline.(14) The samples were joint called with >15,000 other samples and added to seqr(15) (https://seqr.broadinstitute.org), an exome/genome analysis software hosted on the cloud platform Terra (https://app.terra.bio).

#### Genome Medicine Service pipeline

27 patients in 9 families were consented for GMS clinical genome sequencing; however, as one family (FAM_4) comprised a quad, the parents were sequenced with each child as two independent trios. Sequencing was performed on an Illumina NovaSeq 6000 machine, with ≥95% of the autosomal genome covered at ≥15x calculated from reads with mapping quality >10 and >85×10^9 bases with Q≥30, after removing duplicate reads and overlapping bases after adaptor and quality trimming. Cross-sample contamination was checked using VerifyBamID and samples with >3% contamination failed QC. Sequencing alignment was performed using the DRAGEN aligner, with ALT-aware mapping and variant calling to improve specificity. Detection of small variants (single nucleotide variants (SNVs) and indels) and CNVs were performed using the DRAGEN small variant caller and DRAGEN CNV respectively. Short tandem repeat expansions were detected using ExpansionHunter (v2.5.6) as part of the DRAGEN software. The DRAGEN software v3.2.22 was used for alignment and variant calling and structural variants were detected using Manta (v1.5).

### Data analysis

Different analysis strategies were applied to the research exome and the GMS genome data (**Table 2**). The research exome adopted the HiPPo strategy and the GMS adopted a panel-based approach.

**Table 2.**
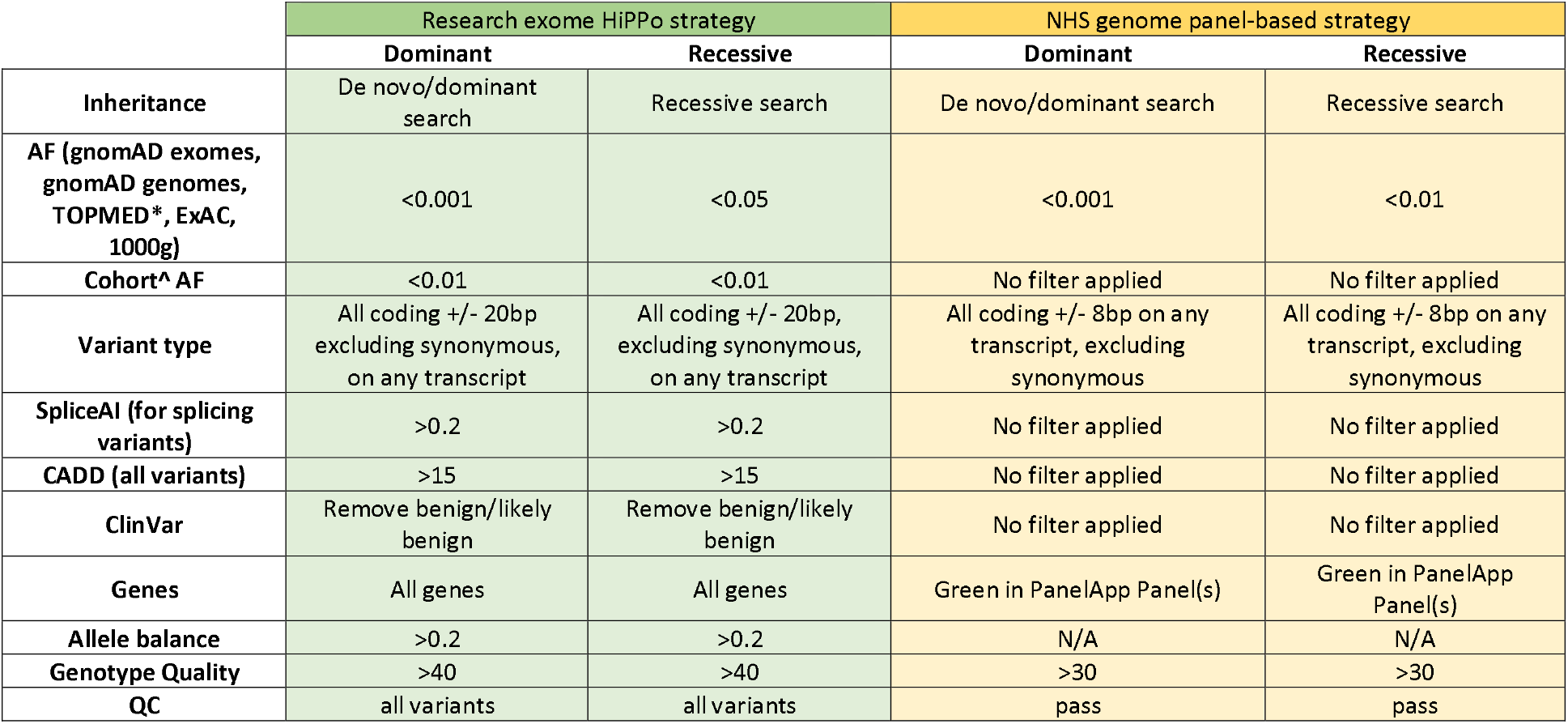
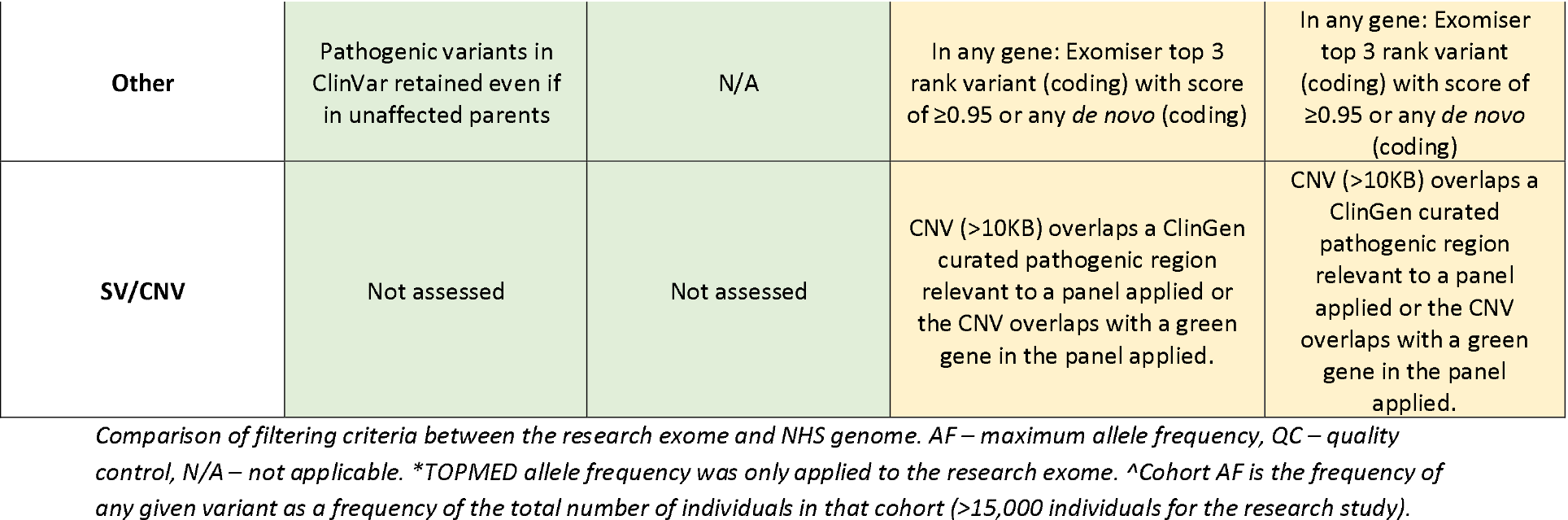
Filtering criteria for the research exome and NHS genome

|  | Research exome HiPPo strategy |  | NHS genome panel-based strategy |  |
| --- | --- | --- | --- | --- |
|  | Dominant | Recessive | Dominant | Recessive |
| Inheritance | De novo/dominant search | Recessive search | De novo/dominant search | Recessive search |
| AF (gnomAD exomes, gnomAD genomes, TOPMED*, ExAC, 1000g) | $<0.001$ | $<0.05$ | $<0.001$ | $<0.01$ |
| Cohort^ AF | $<0.01$ | $<0.01$ | No filter applied | No filter applied |
| Variant type | All coding +/- 20bp excluding synonymous, on any transcript | All coding +/- 20bp, excluding synonymous, on any transcript | All coding +/- 8bp on any transcript, excluding synonymous | All coding +/- 8bp on any transcript, excluding synonymous |
| SpliceAI (for splicing variants) | $>0.2$ | $>0.2$ | No filter applied | No filter applied |
| CADD (all variants) | $>15$ | $>15$ | No filter applied | No filter applied |
| ClinVar | Remove benign/likely benign | Remove benign/likely benign | No filter applied | No filter applied |
| Genes | All genes | All genes | Green in PanelApp Panel(s) | Green in PanelApp Panel(s) |
| Allele balance | $>0.2$ | $>0.2$ | N/A | N/A |
| Genotype Quality | $>40$ | $>40$ | $>30$ | $>30$ |
| QC | all variants | all variants | pass | pass |
| Other | Pathogenic variants in ClinVar retained even if in unaffected parents | N/A | In any gene: Exomiser top 3 rank variant (coding) with score of $\geq 0.95$ or any *de novo* (coding) | In any gene: Exomiser top 3 rank variant (coding) with score of $\geq 0.95$ or any *de novo* (coding) |
| SV/CNV | Not assessed | Not assessed | CNV (>10KB) overlaps a ClinGen curated pathogenic region relevant to a panel applied or the CNV overlaps with a green gene in the panel applied. | CNV (>10KB) overlaps a ClinGen curated pathogenic region relevant to a panel applied or the CNV overlaps with a green gene in the panel applied. |

*Comparison of filtering criteria between the research exome and NHS genome. AF – maximum allele frequency, QC – quality control, N/A – not applicable. \*TOPMED allele frequency was only applied to the research exome. ^Cohort AF is the frequency of any given variant as a frequency of the total number of individuals in that cohort (>15,000 individuals for the research study).*

#### Research exome analysis

For the research exome, each family was analysed as a unit to utilise segregation data. We applied the same *de novo*/dominant and recessive filtering strategies to all families, applying a gene-agnostic filtering strategy by selecting variants with the highest pathogenic potential (HiPPo) using allele frequency, *in silico* prediction scores, and ClinVar (**Table 2**). We later restricted the HiPPo strategy to GenCC(16) genes with a definitive or strong disease association.

#### Reporting on exome variants

Variants remaining following HiPPo filtering were reviewed in seqr(15) using a wealth of in-built annotations. Variants that did not meet any of the below exclusion criteria were considered ‘reportable’ and returned to the referring clinician following application of ACMG-AMP guidelines(17), and any novel discoveries were discussed regarding submission to the Matchmaker Exchange (MME).(18-20)

#### Exclusion criteria

1. The variant was heterozygous in a known autosomal recessive disease gene and no second hit (coding or non-coding) was identified
2. The variant was found in a disease gene and was not associated with the phenotype presented by the patient, as assessed using OMIM(21), GenCC(16) and the medical literature and the variant is not likely pathogenic/pathogenic in ClinVar(12)
3. The variant is in a known disease gene but that gene is poorly expressed as indicated in GTEx(22) in the tissue relevant to the patient’s phenotype or in an exon of the gene with poor expression as determined by the per base expression metric, pext(23)
4. The variant was in a novel gene (currently unassociated with disease) and
  a. the gene is poorly expressed in the relevant disease tissue as indicated in GTEx(22) OR
  b. the gene is explicitly **not** involved in the relevant biological pathway as evidenced in Monarch(24)
5. A predicted loss-of-function (LoF) variant as called by Variant Effect Predictor(25) that was deemed as ‘not LoF’ or ‘likely not LoF’ after application of LoF manual curation guidance as recommended by Karczewski et al.(26)
6. The variant appeared artefactual upon visualisation of the read data in Integrative Genomics Viewer (IGV).(27)

#### Taking novel exome candidates forward

Where the referring clinician agreed, candidate variants in novel genes were submitted by the researcher to MME, sharing anonymised genotype and phenotype data. Any potential matches were discussed in detail with the patient’s clinician and explicit consent was obtained from the participants prior to joining case series.

#### GMS clinical genome analysis pipeline

Variants called through the GMS pipeline were prefiltered on mode of inheritance, quality, and allele frequency. These variants were then restricted to ‘green’ genes on the pre-selected PanelApp(4) gene panels for review (**Table 2**). A complementary gene agnostic filter was also applied to the data, which included all *de novo* variants and Exomiser(6) top 3 ranked coding variants (of any quality). Variants passing filtering were returned to the Wessex Regional Genetics Laboratory for reporting.

#### Reporting of GMS genome variants

GMS variant classification was carried out according to the ACMG/AMP guidelines with ACGS(28) modifications. This included as assessment of the gene-phenotype match based on the HPO(29) terms supplied. Variants in genes with no known disease association (determined using OMIM(21), HGMDPro, ClinGen(30) and PanelApp(4)) were discounted and not assessed. Classified variants were reported according to standard ACMG/AMP guidelines: i.e pathogenic and likely pathogenic variants were always reported, variants of uncertain significance (VUSes) were only reported if there was significant evidence for pathogenicity and/or with the prior agreement of the clinician following a multidisciplinary team discussion (typically via email).

### Comparison of two approaches

We compared the diagnostic yield and the number of variants requiring assessment after variant filtering for both the research exome HiPPo approach and GMS clinical genome panel-based approach. Specifically, we counted the number of variants passing HiPPo filtering criteria in the research exome study and compared these with the number of Tier 1 and 2 variants for the same patients’ GMS genome results, in addition to the ‘gene agnostic’ variants (de novo and Exomiser(6) Top 3 hits) as provided in an anonymised spreadsheet by the Wessex Regional Genetics Laboratory. We omitted CNVs since these were not assessed in the exome data and no diagnoses were made from structural variants in the GMS clinical genome data. We then compared the variants reported from the research exome with the variants interpreted and reported by the NHS on the patient’s GMS genome report. For the GMS, the reporting threshold is high with novel genes and nearly all VUSes not reported. Therefore, to test the efficiency of the methods applied, we calculated the diagnostic rate per number of variants assessed across the cohort.

## Results

27 individuals from 9 families were consented for a GMS clinical genome on the NHS, with 8 families having a report returned. At the time of writing, family FAM_7 is still awaiting genome results, a year after consent was obtained due to requirements for a new maternal blood sample.

26 individuals from the same 9 families underwent exome sequencing at the Broad Institute Center for Mendelian Genomics. Therefore, there were a total of 24 individuals in 8 families who completed parallel research exome and GMS clinical genome sequencing.

### GMS clinical genome analysis strategy

In the 8 families who underwent GMS clinical genome sequencing, a total of 77 single nucleotide and indel variants, were returned for analysis as ‘Tiered variants’ including the gene agnostic variants (Exomiser and de novos). A further 108 CNVs passed filtering. 5 variants in total from 4 patients were included on the final reports issued by the NHS: two diagnoses, one variant of uncertain significance, and compound heterozygous variants (pathogenic and VUS); all five reported variants were also identified by HiPPo in the research exome (**Table 3**).

**Table 3.**
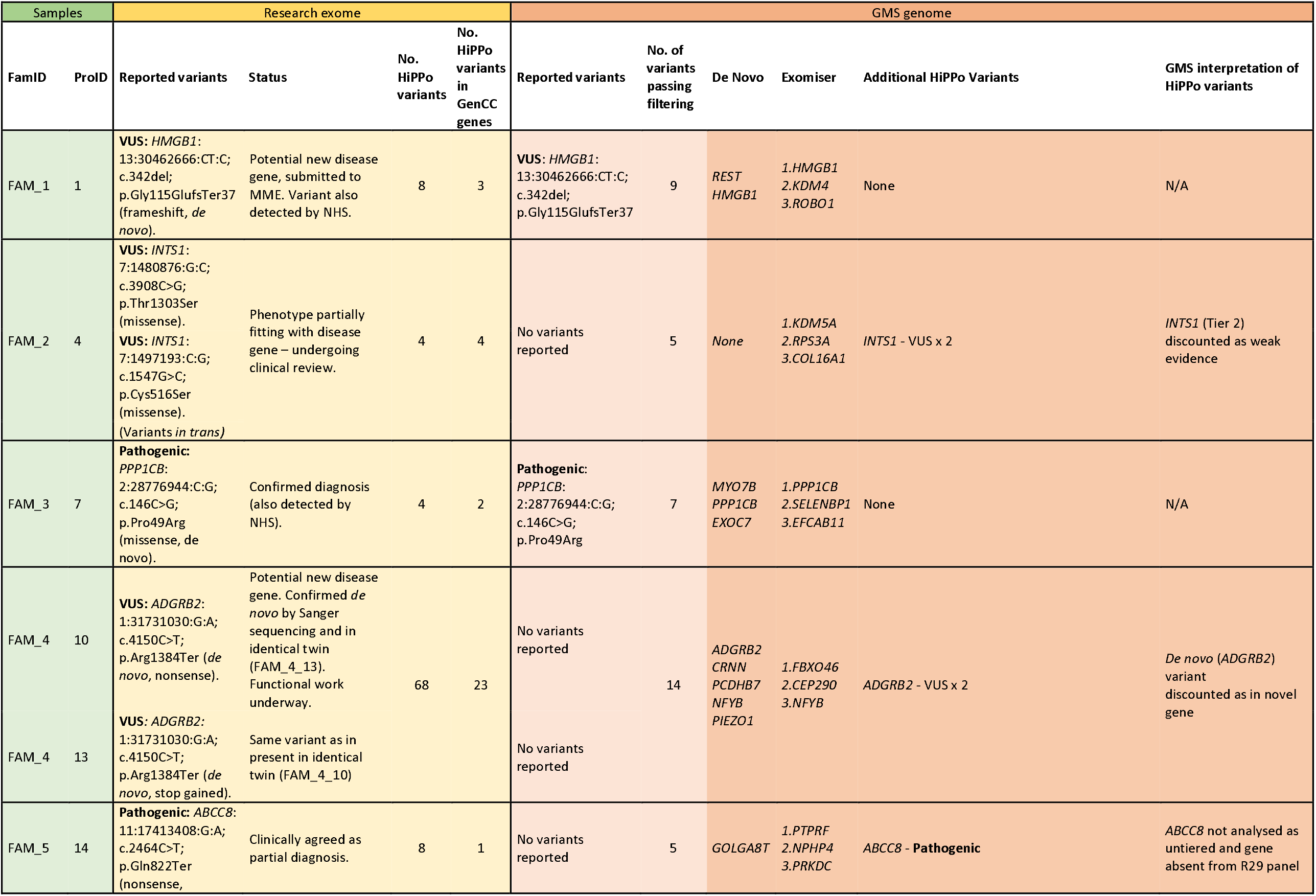
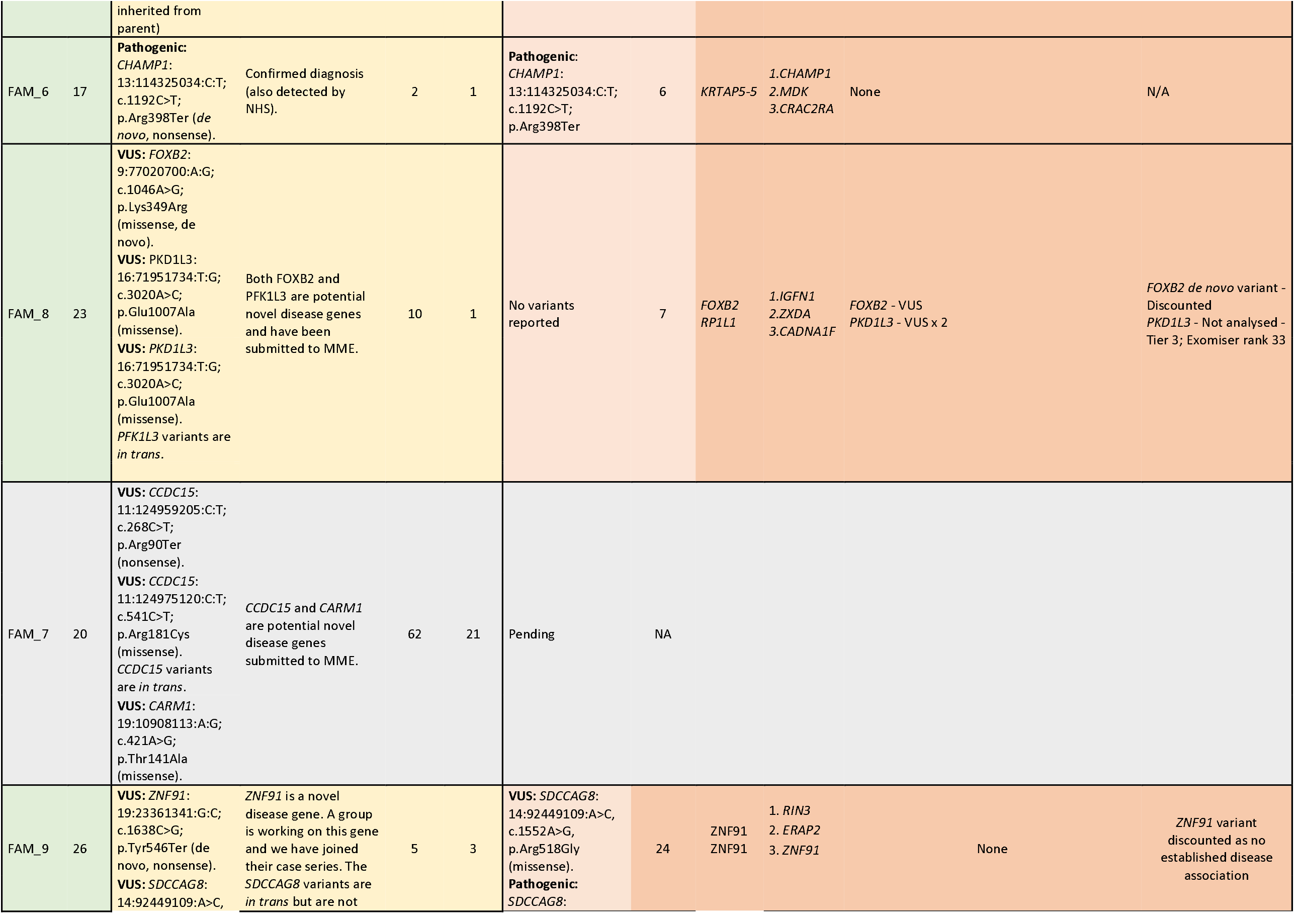
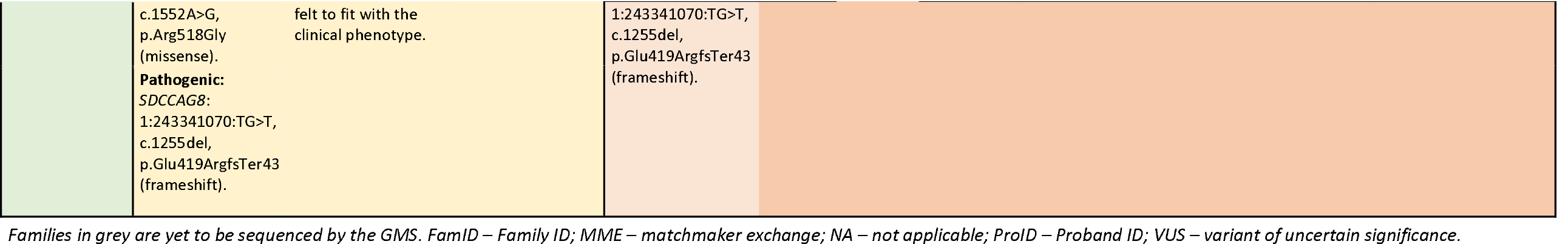
Comparison of variants reported by the research exome sequencing study vs the GMS genome sequencing

### Research exome HiPPo strategy

HiPPo identified a total of 174 variants (**Supplementary Data Table 2**) from 9 families (9 trios) passing filtering criteria as outlined in **Table 2**. When restricting HiPPo to the 8 families who also underwent GMS genome sequencing, HiPPo identified 109 variants. However, one family, FAM_4, comprising a mother, father and monozygotic twins was sequenced as a trio (father, twin A and twin B) in the research exome study as there was insufficient maternal DNA. For the genome performed through the GMS, there was available maternal DNA and thus the twin daughters were sequenced as separate trios, with the parents sequenced twice in accordance with GMS policy. This meant more variants were identified in the research exome than the GMS genome (68 vs 11 respectively) for this family, given that no maternal DNA was available for segregation analysis in the exome.

Of the 174 variants identified by HiPPo across the 9 families, 59 variants were in genes reported as strong evidence for disease association as classified by GenCC.

In addition to the 2 pathogenic variants identified by the GMS and deemed causal, HiPPo identified a further pathogenic variant in a known disease gene (*ABCC8*), representing a partial diagnosis that was filtered out by the GMS strategy due to not being on the chosen gene panel. HiPPo also identified compound heterozygous variants in a known disease gene, *INTS1* in participant FAM_2_4 which is known to cause an autosomal recessive neurodevelopmental disorder with cataracts, poor growth, and dysmorphic facies (MIM: 618571). These variants were discounted by the GMS as weak VUSes with limited evidence but remain under review by the clinical team.

HiPPo detected a further 8 VUSes in 7 novel (currently unassociated with disease) genes, in addition to the same compound heterozygous variants in *SDCCAG8* and the VUS in *HMGB1* reported by the GMS (**Table 3**). In total, the research exome identified 174 variants using HiPPo of which 59/174 (33.9%) were in GenCC disease genes. After application of exclusion criteria to all HiPPo variants, independent of GenCC disease status, a total of 17 variants from the research exome were curated against ACMG/AMP criteria and returned as shown in **Table 4**.

**Table 4.**
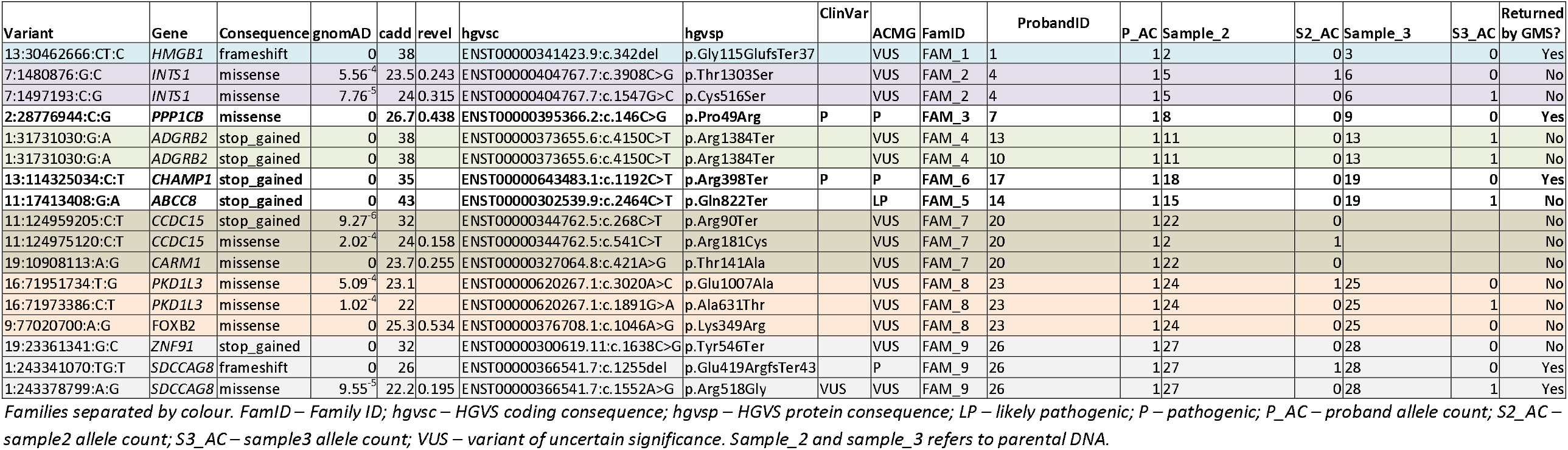
Details of 17 variants reported by the research exome study meeting prioritisation criteria

### Comparison between exome study and GMS clinical genome results

On average, more HPO terms were recorded in the research exome study compared to the GMS genome (**Table 1** and **Figure 4**) although this was not statistically significant (p-value = 0.1, Wilcox signed rank test).

**Figure 4.**
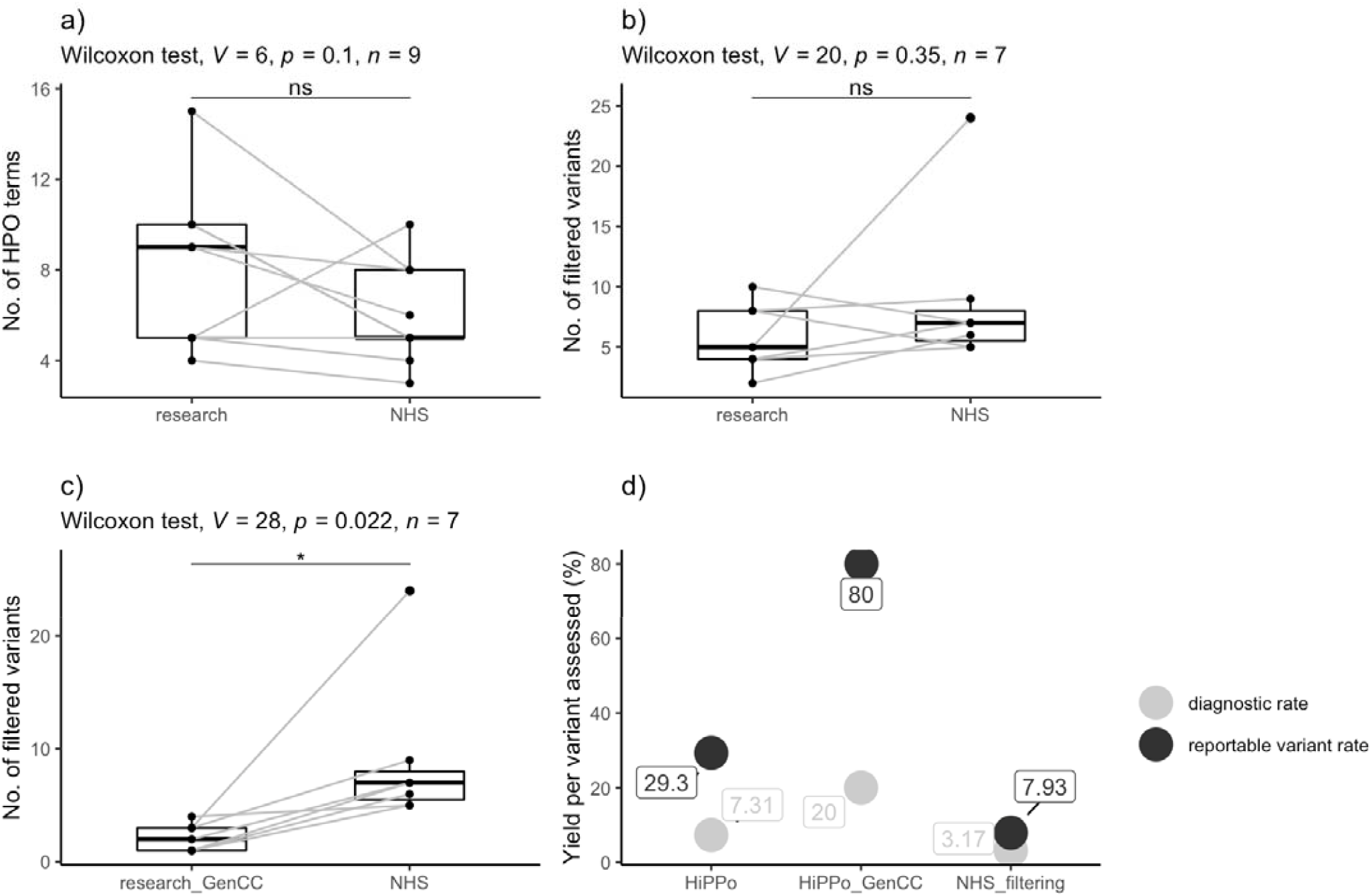
Comparison of results between the research exome and clinical genome (NHS) sequencing (a) Number of HPO terms recorded between the exome and genome studies. (b) Number of variants assessed by the NHS reporting laboratory following GMS genome sequencing versus number of variants passing HiPPo filtering (in any gene) in the exome study. (c) Number of variants assessed by the NHS reporting laboratory following GMS genome sequencing versus the number of filtered HiPPo variants in GenCC disease genes assessed by the exome study. (d) Plot showing the diagnostic rate per variant assessed and the reported variant rate per variant assessed for the HiPPo research approach, HiPPo restricted to GenCC disease genes approach, and the GMS panel-based filtering strategy.

When comparing the 8 families who underwent parallel research exome and GMS clinical genome sequencing, we removed one family (FAM_4) from analysis as the mother was not sequenced in the research study but was sequenced by the GMS. There was no statistical difference between the number of variants (excluding CNVs) assessed by the GMS panel-based strategy and the HiPPo method (p-value = 0.35, Wilcoxon signed rank test), although HiPPo identified more reportable variants (**Table 4**), of which 9 variants in 7 unique genes have been taken forward as candidates to MME. Five of these variants were identified but discounted by the GMS as disease gene discovery is outside of the remit of clinical reporting. However, when restricting the HiPPo analysis to GenCC strong and definitive genes, there was a statistical difference between groups (p-value = 0.022, Wilcoxon signed rank test), with the research study assessing fewer variants overall (**Figure 4**) yet still identifying an additional pathogenic variant in *ABCC8* that did not pass filtering thresholds by the GMS.

The efficiency of the relative analytical methods varied between the groups. The diagnostic rate per number of variants assessed was higher for the HiPPo approach applied to the research exome (3/41 [7.31%]) compared with the panel-based GMS strategy (2/63 [3.17%]). When limiting HiPPo analysis to GenCC disease genes, the diagnostic rate per variant assessed improved further to 3/15 (20%) (**Figure 4**). The reportable variant rate per number of variants assessed was higher for the HiPPo approach when limited to GenCC disease genes (12/15 [80.0%]) compared with the panel-based GMS strategy (5/63 [7.93%]).

## Discussion

Genome sequencing is available as a clinical test on the NHS through the GMS. Following sequencing, data are filtered by a pre-selected gene panel chosen by the referring clinician, in addition to CNVs overlapping the panel applied, *de novo* variants, and Exomiser(6) top 3 ranked variants. This predominantly ‘panel-based’ approach attempts to minimise noise and efficiently identify pathogenic variants in disease-relevant genes.

However, panel-based strategies are not without limitations. PanelApp(4) is open-source but gene reviews and updates of the approved gene content relies on volunteer efforts and comes with a significant lag time. Panels represent a snapshot in time and their application is contingent on clinicians selecting the optimal gene panel(s) with variable levels of genetics training. This is particularly problematic for clinicians in non-genetics specialties lacking adequate familiarity with gene panel selection. If the “wrong” panel is chosen, pathogenic variants can easily be missed. With only 20% of rare disease patients receiving a diagnosis through the 100,000 Genomes Project(1) (the precursor to the UK’s GMS), there is clear need to investigate variants beyond a limited gene list but without significantly increasing the number of variants for review.

This study compares the GMS’ data analysis strategy using genome sequencing to a gene-agnostic HiPPo approach targeting variants with high pathogenic potential as applied to exome sequencing in a research setting. 24 individuals from 8 families underwent parallel clinical genome and research exome sequencing, providing an opportunity to compare these approaches. With many factors influencing timescales between the research and NHS studies, the fairest comparison of efficiency of the two approaches was the number of variants that required review following filtering and the corresponding diagnostic rates. On average the research exome study reviewed fewer variants than the GMS yet identified more diagnostic variants, although this was not statistically significant (p-value = 0.035). The number of reportable variants per variant assessed was higher for HiPPo (29.3%) versus the GMS (7.9%), however the threshold for what constituted a reportable variant differed between the research exome and the GMS genome strategies. The research exome reported variants that would not be reportable in the current NHS setting, although it is worth noting that some international diagnostic labs do report variants in novel genes. However, when restricting the exome HiPPo filtering approach to GenCC disease genes (genes strongly associated with disease that would be reportable in the NHS setting), statistically fewer variants required assessment when compared with the GMS’ panel-based approach (Wilcoxon signed rank p-value = 0.022). Despite this, more pathogenic variants were identified; including a pathogenic variant in *ABCC8* representing a partial diagnosis which was missed by the GMS as it was not on the selected gene panel. For the 8 families undergoing parallel exome and genome sequencing, the GenCC disease gene HiPPo analysis strategy identified 15 variants that required further assessment, compared with 41 variants for the GMS approach. Overall, the diagnostic rate per number of variants assessed between the GenCC disease gene HiPPo analysis and the GMS’ panel-based approach was 3/15 [20%] vs 2/63 [3%] respectively. There is therefore a strong argument that genotype-to-phenotype methods, focused on variants with high pathogenic potential in known disease genes could prove more effective and less resource-intensive than panel-based approaches, despite covering a wider range of the genome. Indeed, in the GMS very few Tier 2 variants are actually reported, meaning that Tier 1 + HiPPo may be prove an efficient alternative strategy and could also be used to prioritise the interpretation of gene agnostic variants and/or determine which should be reported and/or taken to multidisciplinary team meetings. There is also a further argument that genome sequencing is not being optimally utilised by the NHS due to resource limitations and that exome sequencing may prove similarly effective; however, this comparison is beyond the scope of this limited study, whereby no pathogenic CNVs were identified and a time-cost-analysis could not be fairly undertaken.

The number of HPO terms did not vary significantly between those selected for the research study versus those submitted by clinicians working in the NHS (p-value = 0.1) (**Table 1**). A recent study by Kingsmore *et al*.(31) showed that more HPO terms may not increase diagnostic yield, but that a more focused list of key terms may support analysis more effectively.

In total, HiPPo identified 3 diagnoses (compared with 2 diagnoses by the GMS) and a further 12 variants of interest in 8 unique genes, of which 5 genes were discounted by the GMS (**Table 4**) as they did not meet the threshold for clinical reporting. In FAM_2_4, HiPPo identified compound heterozygous variants in *INTS1* (7:1480876G:C and 7:1497193:C:G), a disease gene associated with an autosomal recessive disorder (MIM: 618571) presenting with cataracts, poor growth, developmental delay, and dysmorphic facies. Whilst FAM_2_4 shares some features with the *INTS1* related syndrome, he does not have cataracts and is large (with his weight tracking along the 99^th^ percentile) opposed to being small. These variants are being reviewed by his clinical team.

In FAM_9_26, both HiPPo and the GMS identified compound heterozygous variants (one pathogenic and one VUS) in *SDCCAG8* (1:243341070:TG:T and 1:243378799:A:G), a disease gene associated with an autosomal recessive retinal-renal ciliopathy (MIM: 615993 and MIM: 613615). These variants have been discussed at length with the clinical team and are not felt to explain the nephrotic phenotype. On renal biopsy, the patient had immature glomerular development diffuse foot process effacement on electron microscopy which is inconsistent with a retinal-renal ciliopathy. Furthermore, there were additional inconsistent features including microcephaly, cerebellopontine hypoplasia and functional asplenia. In the same individual, we identified a *de novo* variant in *ZNF91*. Through MME, we are collaborating with a group performing functional studies on this gene, whereby they also have a patient with microcephaly and nephrotic syndrome.

In total, we submitted 9 variants in 7 novel genes to MME from the exome study, which is beyond the remit of the NHS diagnostic capacity. We had no matches for *CCDC15, CARM1, PKD1L3* and *FOXB2*. In addition to *ZNF91* (as described above in FAM_9_26), we matched with collaborators working on *HMGB1* (*de novo* variant found in FAM_1_1) and *ADGRB2* (*de novo* variant found in monozygotic twin sisters FAM_4_13 and FAM_4_10). In 2021, a paper was published on *HMGB1* predicted loss-of-function variants in 6 patients.(32) Common features included developmental delay, language delay, microcephaly, obesity and dysmorphic features, some of which overlap with FAM_1_1. This variant has been returned to the patient’s clinician and we have put them directly in touch with the authors of the 2021 paper for an ongoing collaboration. Whilst the *HMGB1* variant was also reported as a VUS through the GMS, there is no time provision for clinicians to consider and follow up any unreported novel candidates. Furthermore, most *de novo* candidates in novel genes are disregarded by the GMS and so are seldom investigated further. That said, anonymised patient data are eventually deposited in the Genomics England Research Environment, meaning that novel variants may be identified and later investigated through research.

In 2017, a paper was published in Human Mutation describing a missense variant in *ADGRB2* in a patient presenting with developmental delay and progressive spastic paraparesis; features shared with identical twins FAM_4_13 and FAM_4_10 harbouring a *de novo* pLoF in the same gene.(33) The authors showed that their specific variant demonstrated gain of function. We have contacted the authors of the paper and are now directly working with them to model our variant *in vitro* and *in vivo*.

### Limitations

This study is small, representing 27 individuals from 9 families, of which 24 participants received parallel exome and genome sequencing. Inevitably a larger study is needed to test the value of gene-agnostic approaches utilising pathogenicity scores compared with gene panel approaches. This is not easily feasible within the NHS, as it is not possible to access an individual patient’s sequencing data through the GMS to test alternative strategies. Therefore, the only way to compare methods was in a study that independently sequenced the same patients.

Data analysed in a research setting is not comparable with data analysed for diagnostic purposes as the threshold for variant follow-up and investigation may differ in a clinical setting, with inconsistency in reporting on novel discoveries.

No pathogenic variants were identified by GMS clinical genome sequencing that were not captured by the research exome, although a larger sample size is needed to test the diagnostic uplift gained from structural variants detected using genome sequencing versus potential missed diagnoses from using panel-based approaches.

## Conclusion

This study compared a gene agnostic filtering strategy called HiPPo as applied to research exome data with a gene panel-based analysis strategy applied to genome sequencing data. Despite HiPPo being pan-exomic, a similar number of variants were assessed per patient to the panel-based strategy of the GMS and more variants of interest were identified; this includes a pathogenic variant in *ACDCC8* and *de novo* variants in 3 novel genes, whereby case series and functional experiments are underway. When restricting HiPPo to GenCC disease genes, statistically fewer variants required assessment to identify the same diagnoses as identified by the GMS (20% vs 3% respectively), representing a greater diagnostic yield per variant assessed. This work suggests that panel-based approaches are limited and that they could be improved by incorporating specific variant prioritisation metrics. Further testing is required to integrate these complementary approaches to optimise the analytical strategy for genome sequencing within the NHS.

## Data Availability

All data produced in the present work are contained in the manuscript

## Supporting information

### Supplementary Data

<u>Supplementary Table 1:</u> DNA concentration and volume available for samples consented for research exome sequencing

<u>Supplementary Table 2:</u> All HiPPo filtered variants following research exome sequencing of 27 individuals in 9 families

